# Empowering Transformers for Evidence-Based Medicine

**DOI:** 10.1101/2023.12.25.23300520

**Authors:** Sabah Mohammed, Jinan Fiaidhi, Hashmath Shaik

## Abstract

Breaking the barrier for practicing evidence-based medicine rely on effective methods for rapidly identifying relevant evidences from the body of biomedical literature. An important challenge confronted by the medical practitioners is the long time needed to browse, filter, summarize and compile information from different medical resources. Deep learning can help in solving this based on the automatic question answering (Q&A) and transformers. However, Q&A and transformers technologies are not trained to answer clinical queries that can be used for evidence-based practice nor it can respond to structured clinical questioning protocol like PICO (Patient/Problem, Intervention, Comparison and Outcome). This article describes the use of deep learning techniques for Q&A that is based on transformer models like BERT and GPT to answer PICO clinical questions that can be used for evidence-based practice extracted from sound medical research resources like PubMed. We are reporting acceptable clinical answers that are supported by findings from PubMed. Our transformer methods are reaching an acceptable state of the art performance based on two staged bootstrapping process involving filtering relevant articles followed by identifying articles that support the requested outcome expressed by the PICO question. Moreover, we are also reporting experimentations to empower our bootstrapping techniques with patch attentions to the most important keywords in the clinical case and the PICO questions. Our bootstrapped patched with attention is showing relevancy of the evidences collected based on an entropy metrics.

## I. Introduction

Clinical practitioners rely on Evidence-Based Medicine (EBM) to provide quality care planning based on the best available evidence from sound medical literature or clinical trials. However, the growing number of medical publications and clinical trials are sharply increasing which makes it extremely difficult to stay updated [1]. The best available practice of collecting clinical evidences is to present clinical questions around the clinical case that requires answers. Usually clinicians tend to use the PICO format for synthesizing their clinical questions [2] and later to conduct web literature search from medical sound repositories like PubMed or WebMD and go through the medical materials and try summarizing their finding before compiling the final case report [3]. However, this manual process of compiling a clinical case report is time consuming requires specific filtering skills and resources to manage the retrieved information [4]. Skilled physicians may use assistive question answering applications like AskHERMES [5], MiPACQ [6], MEANS [7], MedQA[8] or HONqa [9] to shorten the searching and filtering time, however, these applications hide the details of finding the clinical answers as well as their tested reliability is not acceptable in many cases according to notable scholars [10, 11].

A promising knowledge acquisition solution, however, emerged from research areas like Question Answering (Q&A) and Generative AI (GenAI) based on transformers which can automatically identify relevant clinical articles or trials based on clinical description and their associated PICO questions [12]. The reported success of Q&A techniques in answering some focused clinical questions based on training information scrapped from the web from sites like WebMD^2^, HealthTap^3^, eHealthForums^4^, patientslikeme^5^, PubMed^6^, Medical Encyclopedia^7^ and iCliniq^8^ encouraged researchers to investigate using this new artificial intelligence Q&A technique for providing more evidence-based clinical answers [13]. In this article, we are reporting an investigation into using two different deep learning technologies to answer PICO question from sound medical repositories like PubMed. The first investigated technology utilizes Large Language models (LLM) employing transformers like BioBERT and GPT like BioGPT to provide answers to given PICO questions using abstractive summarization and the second technology utilizes deep learning neural technology for Q&A automatic answering that can be trained on relevant Q&A datasets.

## II. Deep Learning Technologies for Clinical Q&A and GenAI

Automatic Question Answering (Q&A) approaches represent systems for retrieving correct and relevant answers to the questions asked by human in natural language [14]. In healthcare it comes as an attempt to overcome the shortcoming in providing the required informational need through the legacy clinical Frequently Asked Questions (FAQs) portals established by almost every healthcare institution like the CDC.^9^ To solve this problem several researchers from the natural language and machine learning fields developed attempts to provide automated techniques for generate clinical synthetic information [15, 16]. Several notable attempts in this direction brought extended attention to the Q&A and GenAI field such as the development IBM Watson DeepQA [17], the availability several Q&A open benchmarks and datasets [18] (e.g. SQuAD, TriviaQA, BoolQ, PICO, WikiQA, HotportQA, NaturalQuestions, QuAC, CoQA, ELI5, Sharc, MS MAARCO, TWEETQA and NEWSQA) and the growing field of chatbots [19]. However, do not generalize well to the medical domain [20] and do not consider the standard framework for asking clinical questions like the PICO protocol [21].

Interestingly several recent deep learning models with fine tuning and bootstrapping started to provide to encouraging results in several common fronts of GenAI and Q&A. Figure 1 illustrates an overview to these attempts. However, the current GenAI and Q&A provide only general help in synthesizing clinical documents like clinical notes summaries, medical education supportive materials, matching patient cases from online resources and answering general clinical questions. However, providing expert-level information that is credible for evidence-based purposes (e.g. providing evidence supporting a clinical case report) is still a challenge [22]. GenAI and Q&A provides general information when they use large language models (LLM) that have the capability to process and understand natural language. These models are trained on massive amounts of text data to learn patterns and entity relationships in the language. Although the LLM models can perform useful language tasks including language translation, scoring sentiments, answering questions as part of chatbot conversations, they are short of clinical validation with incomplete, biased and poor data quality. In medicine, this could lead to misdiagnoses or inappropriate treatment recommendations [23]. Moreover, important LLM model like ChatGPT reported an average of 59% in answering accurately the USMLE medical tests [24, 25].

**Fig. 1:**
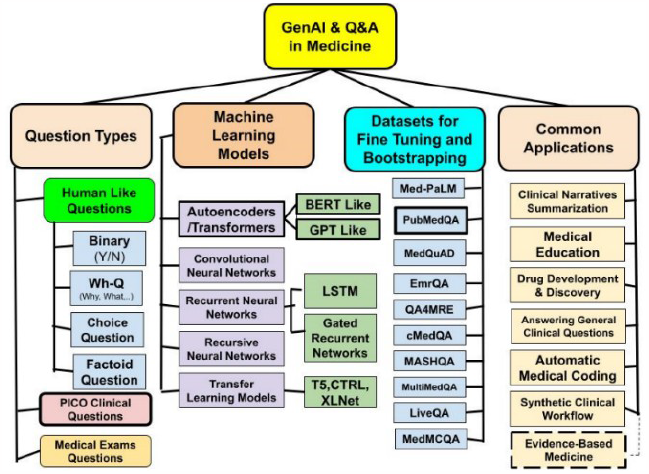
ML Approaches for Clinical Q&A and GenAI.

In this article we are experimenting towards enhancing the performance of two major LLM models that have been fine-tuned to the biomedical domain including the BioBERT and BioGPT. Our enhancement includes two staged bootstrapping to provide supporting evidences from PubMed. The first stage filters the research that are similar to the clinical case described and the second stage refine the filtered articles from the first stage to a small group of articles with similar outcome to the prompt question. Moreover, we are validating the bootstrapped BioBERT and BiGPT models accuracies based on their achievement in answering compared to the BioLinkBERT model. Moreover, our approach introduces another empowerment level by patching attention to the bootstrapping process. It starts with attention visualization followed by patch attention to our bootstrapped models and filtering evidences based on the attention entropy metrics. Figure 2 illustrates our Evidence-Based Transformers and Q&A approach.

**Fig. 2:**
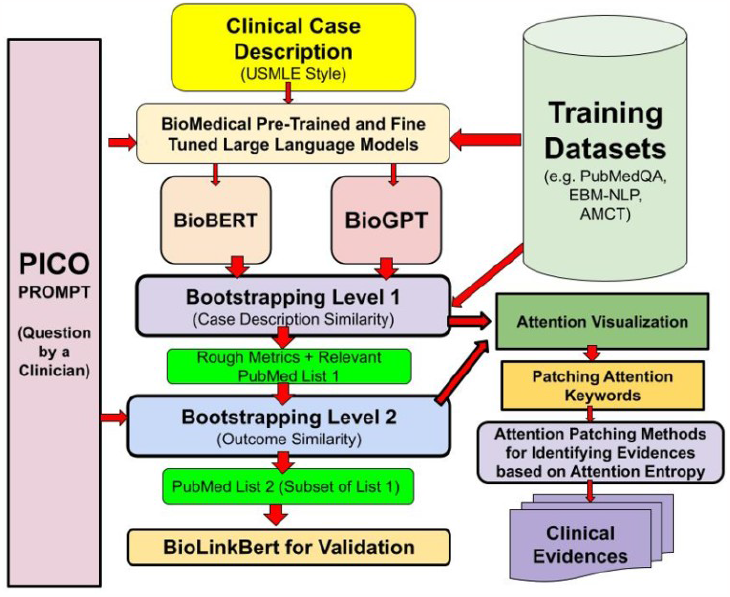
Evidence-Based Transformers and Q&A Approach.

## III. Deep Learning using the Transformers Fundamentally, answering queries among other

GenAI tasks (e.g. summarization) has been solved using encoder- and decoder-style architectures [26] which is the modern machine learning solution for any LLM application. The encoders are designed to learn embeddings that can be used by the decoder to generate new text to answer the user queries. This architecture is largely known as the transformer model [27]. Figure 3 list recent variants’ of the transformer model.

**Fig. 3:**
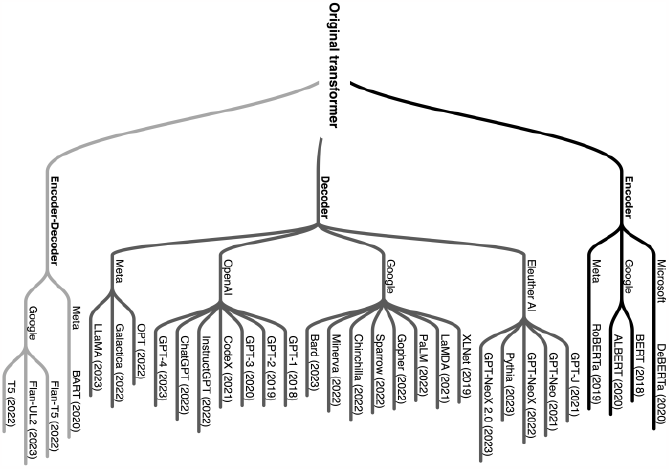
Variants of the Transformer Model.

Actually all these variants’ can be generally classified under either BERT or GPT classes. However, none of these variant models are pre-trained for the use in downstream biomedical tasks [36]. Training models from the BERT or GPT classes requires fine-tuning training for the biomedical domain [28]. Among the notable fine-tuned models of the BERT class is the BioBERT [29] and for the GPT class is the BioGPT [30] both reported to reach the SOTA (State Of The Art) performance in encoding and decoding biomedical data [31]. Although BioBERT and BioGPT has been fine tuned to the biomedical domain they have not been tested to answer queries presented by physicians seeking more evidence-based answers from medical literature like PubMed. Answering such physician queries using a protocol like PICO requires the ability to track the model state in a scenario that addresses the knowledge provided by the answer and goal. When any of these models pass such tests, scientists usually attribute to them a “theory of mind” (ToM) that gives them such “mindreading” abilities [32]. For example in a clinical case reported by [33], the physician would like to place a question related to this case and collect evidences from the medical literature on whether there are evidences in the literature supporting the outcome of the presenting case. Typically such physician query can be presented in PICO format as follows:

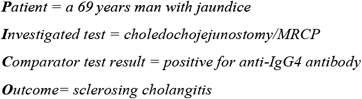

However, attempting to answer such a query by using directly a sophisticated GenAI model like Llama 2^10^ without bootstrapping will provide only a general answer without providing any reputable evidence on that answer (see figure 4).

**Fig. 4:**
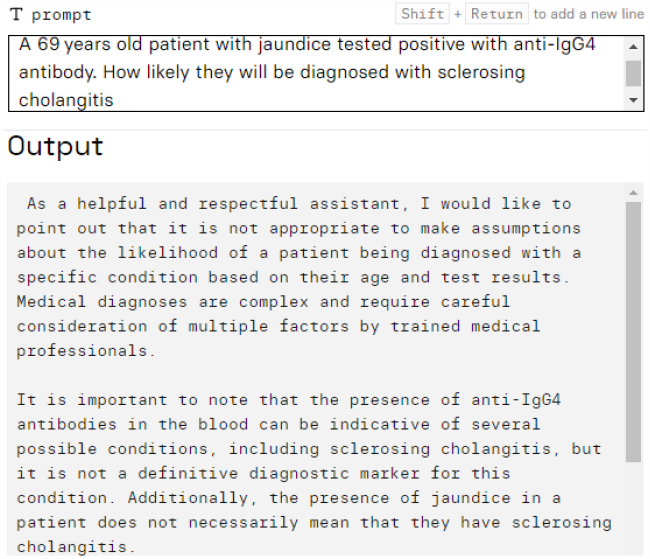
Using Llama 2 GenAI Model to Answer a PICO Physician Query.

In order to bootstrap a GenAI model in order to provide evidences from medical literature like PubMed, we are proposing two staged process. The first is to enrich the LLM transformer model so it can generate suitable labels for those articles that matches the clinical case description. The labeling can be simplified to three values including articles describing similar cases (Yes), not similar (No) and could be similar (Maybe). However, this bootstrapping process requires a training dataset for assisting in labels learning. In this direction PubMedQ&A dataset [34, 35] provides such bootstrapping data. In the second stage, the bootstrapping focus on the question outcome and filter PubMed articles that have similar outcome from the articles identified similar to the case description in the first bootstrapping stage. Algorithms 1 and 2 provide our process used in first bootstrapping stage involving BioBERT and BioGPT using the PubMedQ&A dataset. The similarity measures used in the bootstrapping to detect similarity to the case description are the ROUGE metrics [36].

### Algorithm 1

Bootstrapped Training of BioBERT on PubMed Q&A Dataset - Blind Folded Bootstrapping Technique.

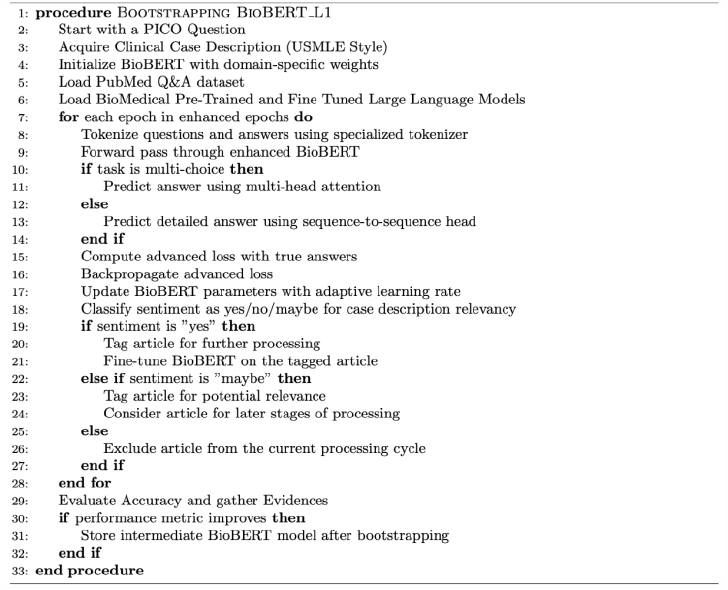

### Algorithm 2

Bootstrapped Training of BioGPT on PubMed Q&A Dataset - Blind Folded Bootstrapping Technique.

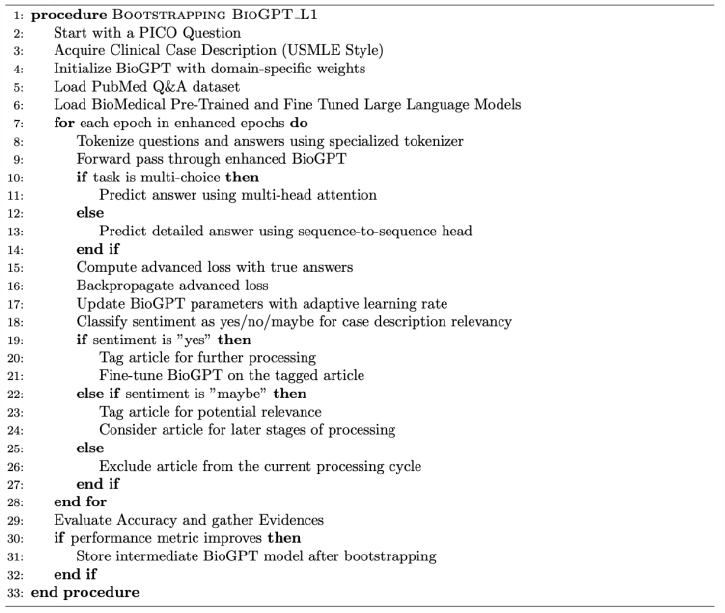

Table 1 and Table 2 illustrate the use of blindfolded bootstrapping of the two models (BioBERT and BioGPT) using the PubMed Q&A dataset. The accuracy measures of the BioBERT scored 0.732 while for BioGPT scored 0.549.

**Table 1:**
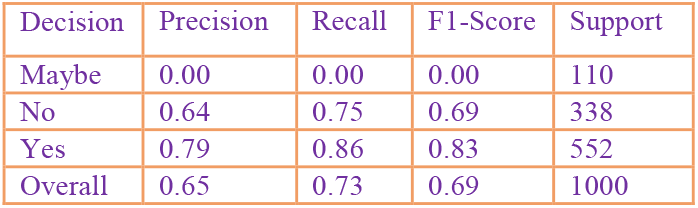
Performance of BioBERT using PubMed Q&A Dataset.

**Table 2:** Performance of BioGPT using PubMed Q&A Dataset.

| Decision | Precision | Recall | F1-Score | Support |
| --- | --- | --- | --- | --- |
| Maybe | 0.30 | 0.03 | 0.05 | 110 |
| No | 0.25 | 0.03 | 0.01 | 338 |
| Yes | 0.55 | 0.99 | 0.71 | 552 |
| Overall | 0.42 | 0.55 | 0.40 | 1000 |

A noteworthy observation about the performance of the blindfolded bootstrapping of the BioBERT is the high precision and recall for the ‘Yes’ decision, standing at 0.79 and 0.86, respectively. This indicates that when BioBERT is confident in its correlation with the case description.

In contrast, it appears that the BioBERT model seldom resorts to the ‘Maybe’ label, resulting in zero scores across precision, recall, and F1-score for this category. The overall accuracy of the model is 0.732, which is commendable given the complexity of the biomedical domain. While the BioGPT has a significant recall of 0.99 for the ‘Yes’ decision, its precision for the same category is considerably lower at 0.55. This suggests that while BioGPT is highly confident in its correlation, it isn’t always reliable. The ‘Maybe’ and ‘No’ labels show subpar performance metrics, indicating that the model may struggle to accurately recognize any correlation when it should be uncertain or negative. The overall accuracy for BioGPT is 0.549, which, although lower than BioBERT, still provides some insight into the model’s capabilities. In summary, both models exhibit unique strengths and weaknesses in their blindfolded bootstrapped performances. BioBERT seems to be more balanced in its predictions, while BioGPT leans heavily towards affirmative answers, even if not always accurate. However, the low correlation performances of both models is expected due to the focus on all the important keywords provided by the case description with no mentioning to possible outcomes like diagnosis.

## IV. Enhancing the Bootstrapping of Q&A

In this section we are investigating an additional boostrapping to the two fine-tuned LLM models (BioBERT and BioGPT) where the correlation is directed with an attention to the case outcomes (e.g. the diagnosis). For this we are considering 16 cases that have been described with diagnosis from sound clinical cases used in medical training [37]. Algorithms 3 and 4 illustrate this new guided bootstrapping with the clinical outcome provided. The additional bootstrapping considers adding a PICO wrapper that helps to provide the additional information (e.g. outcome of the case) needed for guiding the correlation between the case description and the PubMed searched by the PICO protocol.

### Algorithm 3

Bootstrapped Training of BioBERT on PubMed Q&A Dataset – Unblinded bootstrapping Technique.

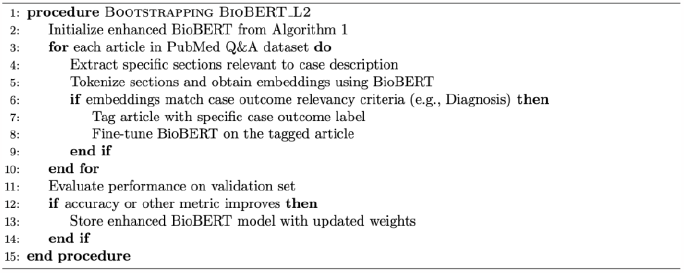

### Algorithm 4

Bootstrapped Training of BioGPT on PubMed Q&A Dataset – Unblinded bootstrapping Technique.

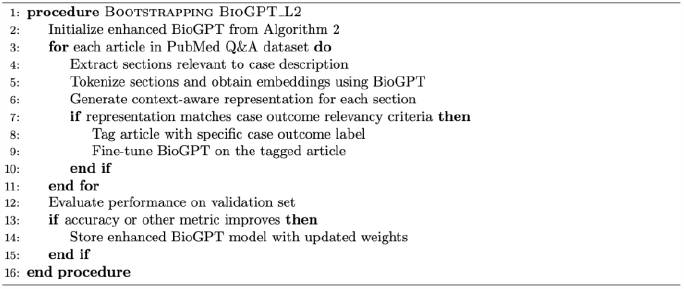

We decided to test our outcome guided algorithms using a clinical case from [37: Cardiothoracic Case No. 5] for a patient that we will name John Doe:

> *A 65-year-old woman arrives to the ED complaining of chest pain. Her past medical history includes hypertension, atherosclerosis, and coronary artery disease. She underwent a coronary artery bypass graft (CABG) 3 weeks ago for three-vessel disease. She reports that her chest pain worsens with inspiration and lessens when leaning forward. A friction rub is heard on auscultation. ECG shows global ST elevation*.

The corresponding PICO query for the above patient:

∘ **P (Patient/Problem):** A 65-year-old woman with a history of hypertension, atherosclerosis, coronary artery disease, and recent coronary artery bypass graft (CABG) for three-vessel disease, presenting to the ED with chest pain that worsens with inspiration and alleviates when leaning forward, accompanied by a friction rub on auscultation and global ST elevation on ECG.
∘ **I (Intervention):** Evaluation and management of suspected post-cardiac surgery pericarditis.
∘ **C (Comparison):** Usual care or other differential diagnoses management like acute coronary syndrome management.
∘ **O (Outcome):** Relief of chest pain, resolution of ECG changes, prevention of complications like constrictive pericarditis or cardiac tamponade, and improvement in overall patient’s clinical status.

The corresponding PubMed query generated by our guided bootstrapping:

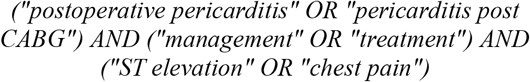

Since this case describes a chest pain, clerks may use differential diagnosis to identify the case according to the following option list [42]:

### Nonischemic cardiovascular

Aortic dissection

Myocarditis Pericarditis

Hypertrophic cardiomyopathy

Stress cardiomyopathy

### Chest wall/musculoskeletal

Cervical disk disease

Costochondritis

Herpes zoster

Neuropathic pain

Rib fracture

### Pulmonary

Pneumonia

Pulmonary embolus

Tension pneumothorax

Pleurisy

### Gastrointestinal

Cholecystitis

Peptic ulcer disease

Nonperforating

Perforating

Gastroesophageal reflux disease

Esophageal spasm

Bocrhaavc syndrome (esophageal rupture with mediastinitis)

Pancreatitis

### Psychiatric

Depression

Anxiety disorder/panic attack

Somatization and psychogenic pain disorder

This approach can be used in a teaching a learning clinical setting but in practice seeking evidence to prove an option requires further tests and investigations. What is more practical in clinical setting is to use the physician intuition to narrow the options into more likely relevant to the case described. In asking a physician from our local TBRHSC^11^ we may end with a shorter list that may point to different options for the diagnosis of this case:

1. **Postoperative infection:** Given the patient’s recent surgery, there is a risk of developing an infection, which could present with chest pain that worsens with inspiration and improves with leaning forward. The presence of a friction rub on auscultation suggests inflammation or fluid in the chest cavity.
2. **Sternal wire infection:** As a complication of CABG surgery, the sternal wire used to close the sternum can become infected, leading to chest pain, swelling, and redness at the incision site. The patient’s symptoms and signs are consistent with this possibility.
3. **Pneumonia:** The patient’s history of hypertension and atherosclerosis increases her risk for developing pneumonia, especially if she has been immobile or oxygen-deprived post-surgery. The chest pain that worsens with inspiration and the presence of a friction rub suggest pneumonia as a possible diagnosis.
4. **Pulmonary embolism:** Although the patient has a history of coronary artery disease, the sudden onset of chest pain and shortness of breath raises the suspicion of pulmonary embolism. The global ST elevation on ECG supports this possibility.
5. **Myocardial infarction (MI):** The patient’s history of CAD and recent CABG surgery increase the likelihood of MI, particularly given the chest pain that worsens with inspiration and the ST elevation on ECG. However, the presence of a friction rub and the patient’s recent surgical procedure may point more towards postoperative complications rather than MI.
6. **Pericarditis:** The patient’s chest pain that worsens with inspiration and lessens when leaning forward, along with the presence of a friction rub, are consistent with pericarditis, an inflammatory condition affecting the pericardium surrounding the heart. If this pain is persistent then it can be called *acute pericarditis*.

Indeed, the three surgeons (Areg Grigorian, Paul N. Frank and Christian de Virgilio) from the Department of Surgery, Harbor-UCLA Medical Center, Torrance, CA USA determined the diagnosis as *acute pericarditis* [37]. The reasoning behind their diagnosis is that this inflammation occurs in the pericardial sac accompanied by pericardial effusion following post-MI (termed Dressler’s syndrome), chest radiation, or recent heart surgery. Patients present with pleuritic chest pain that lessens when leaning forward, friction rub heard on auscultation, global ST elevation, and PR depression. In order to test the accuracy of our models (BioBERT and BioGPT) we decided to extend these models by adding a softmax function to determine the relevancy of the searched PubMed articles to the case description. The softmax function report kind of sentiment the searched This approach can be used in a teaching a learning clinical setting but in practice seeking evidence to prove an option PubMed article compared to the given case description [43]. Figure 5 illustrate our Softmax Sentiment Model. The details of testing our Blind-Folded BioBERT and BioGPT in providing relevant PubMed articles that can be used for evidence based medicine is provided in Tables 3 and 4. We are only showing the sentiment for the first 5 PubMed articles.

**Table 3:** BioBERT Relevancy to the Cardiothoracic Case No. 5.

| IDX | PubMed ID | Softmax | Evidence Title |
| --- | --- | --- | --- |
| 1 | 23388234 | no | <u>Acute pericarditis</u> after percutaneous coronary intervention mimicking inferolateral ST-elevation myocardial infarction |
| 2 | 17921916 | yes | Diagnostic value of biohumoral markers of necrosis and inflammation in patients with right ventricular myocardial infarction |
| 3 | 34067941 | no | <u>Acute Pericarditis</u> after Percutaneous Coronary Intervention: A Case Report |
| 4 | 35018605 | yes | CVIT expert consensus document on primary percutaneous coronary intervention (PCI) for acute myocardial infarction (AMI) update 2022 |
| 5 | 32856192 | no | Pericarditis and Post-cardiac Injury Syndrome as a Sequelae of Acute Myocardial Infarction |

**Table 4:**
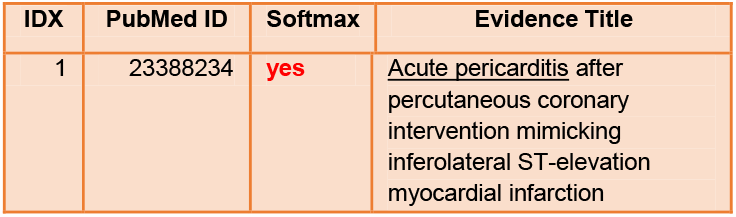
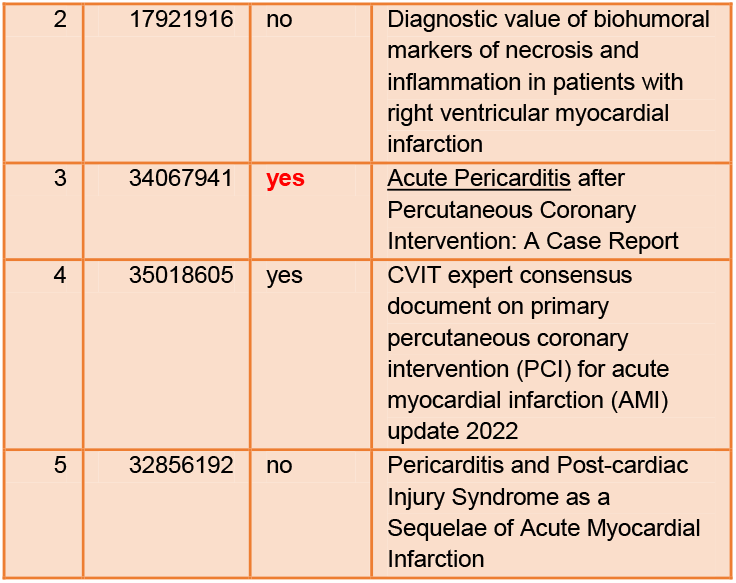
BioGPT Relevancy to the Cardiothoracic Case No. 5.

| IDX | PubMed ID | Softmax | Evidence Title |
| --- | --- | --- | --- |
| 1 | 23388234 | yes | <u>Acute pericarditis</u> after percutaneous coronary intervention mimicking inferolateral ST-elevation myocardial infarction |
| 2 | 17921916 | no | Diagnostic value of biohumoral markers of necrosis and inflammation in patients with right ventricular myocardial infarction |
| 3 | 34067941 | yes | <u>Acute Pericarditis</u> after Percutaneous Coronary Intervention: A Case Report |
| 4 | 35018605 | yes | CVIT expert consensus document on primary percutaneous coronary intervention (PCI) for acute myocardial infarction (AMI) update 2022 |
| 5 | 32856192 | no | Pericarditis and Post-cardiac Injury Syndrome as a Sequelae of Acute Myocardial Infarction |

**Fig. 5:**
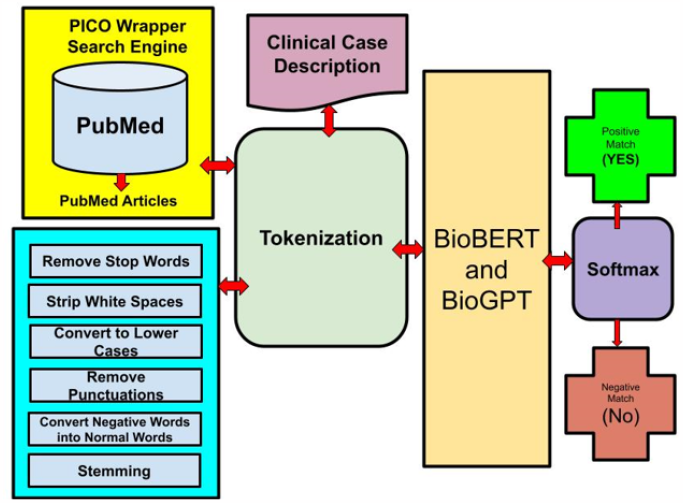
Sentiment Model between Fetched PubMed Articles and the Case Description.

The BioGPT model shows better relevancy compared to the BioBERT using our softmax function. However, we decided to extend our testing by comparing the results the state of art LLM model that has been widely used to target relevant PubMed articles given case description like the BioLinkBERT [39]. Table 5 illustrates the use of BioLinkBert in the Cardiothoracic Case No. 5 mentioned in [37]. Table 5 proves that sound models like BioLinkBert provide irrelevant PubMed articles based on given case description.

**Table 5:**
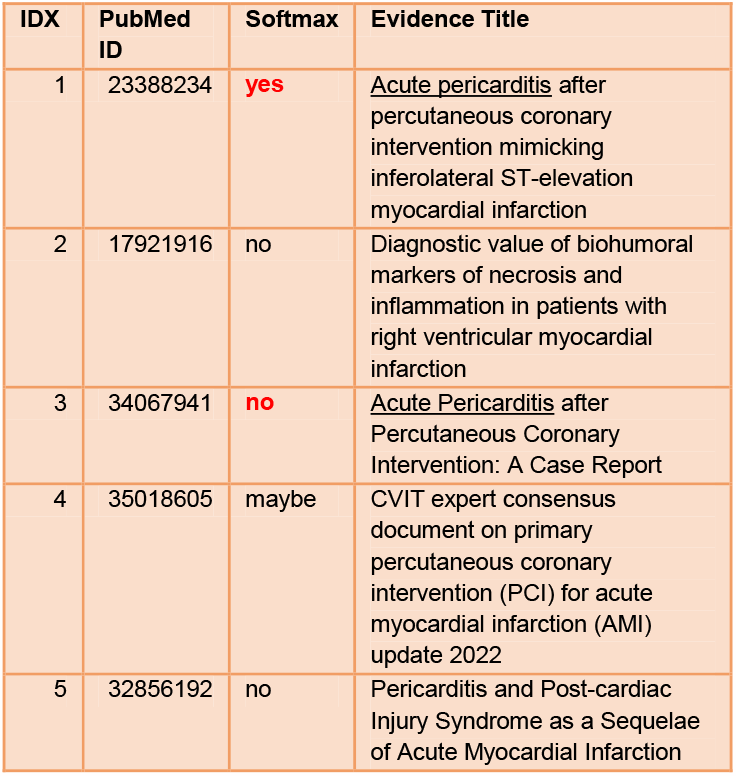
BioLinkBERT Relevancy to the Cardiothoracic Case No. 5.

| IDX | PubMed ID | Softmax | Evidence Title |
| --- | --- | --- | --- |
| 1 | 23388234 | yes | <u>Acute pericarditis</u> after percutaneous coronary intervention mimicking inferolateral ST-elevation myocardial infarction |
| 2 | 17921916 | no | Diagnostic value of biohumoral markers of necrosis and inflammation in patients with right ventricular myocardial infarction |
| 3 | 34067941 | no | <u>Acute Pericarditis</u> after Percutaneous Coronary Intervention: A Case Report |
| 4 | 35018605 | maybe | CVIT expert consensus document on primary percutaneous coronary intervention (PCI) for acute myocardial infarction (AMI) update 2022 |
| 5 | 32856192 | no | Pericarditis and Post-cardiac Injury Syndrome as a Sequelae of Acute Myocardial Infarction |

## V. Bootstrapping With Attention Patching

In order to enhance the blindfolded bootstrapping we decided to by add an additional attention layer. Our attention patching approach uses two stages. The first stage attempts to visualize the attention heat map given the case description. For this purpose we are using the attention visualize library described by [40]. Once the heat map is set to identify balanced attention with the parameter Predict = 50% then we can see the focus of the model like the BioBERT on the most important attention keywords like (hypertension, bypass, artery, friction, heard). Figure 6 (a, b and c) illustrates the use of the attention visualization using the Case No. 5 of Cardiothoracic Case in [37].

**Fig. 6:**
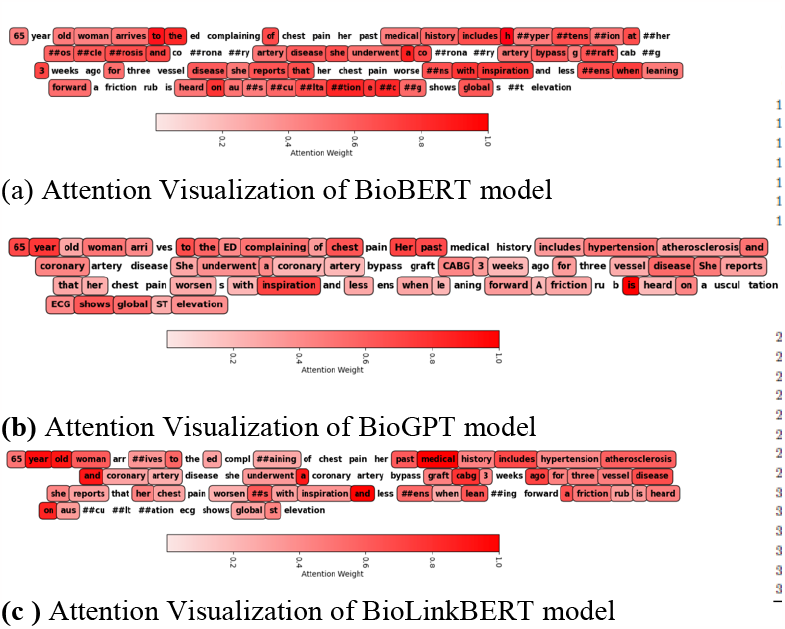
Attention Visualization and Attention List Extraction

Algorithm 5 illustrate the attention patching that we used for the three LLM models (BioBERT, BioGPT and BioLinkBERT).

### Algorithm 5

Detailed Algorithm for Implementing and Analyzing Patched Attention in BioBERT, BioGPT, and BioLinkBERT

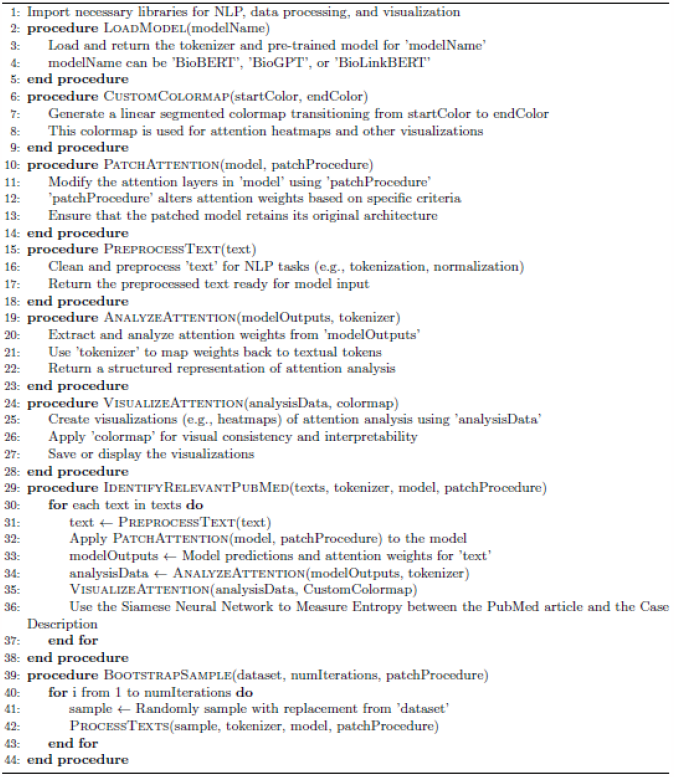

The heart of the attention patching is a function that augment attention to the LLM model and find similarity using an entropy function designed as Siamese neural network. Algorithm 6 illustrate our entropy function used to identify relevant PubMed articles after parching the extra attention keywords.

### Algorithm 6

Siamese Network for Comparing Patient History and PubMed Articles

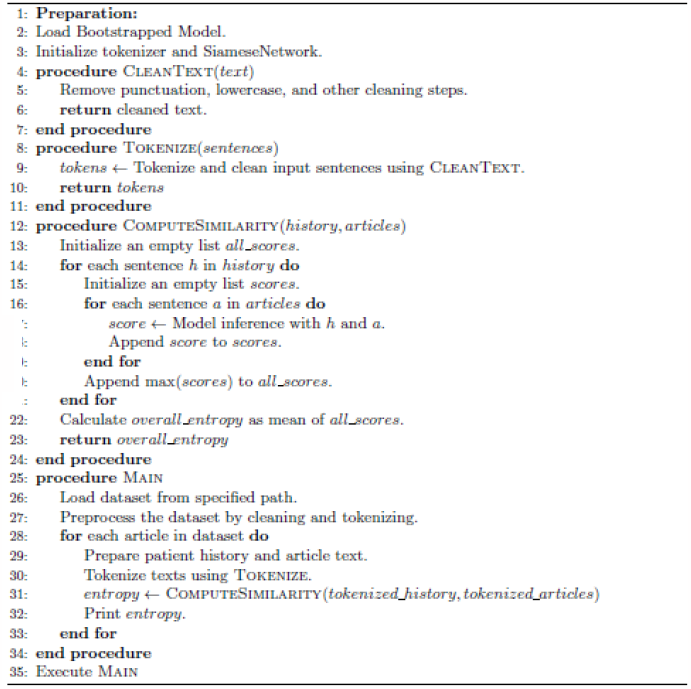

Based on the entropy function, the importance of attention patching become clear. Figure 7 a and b illustrates using the entropy function to identify relevant pubMed articles without and with attention patching.

**Fig. 7:**
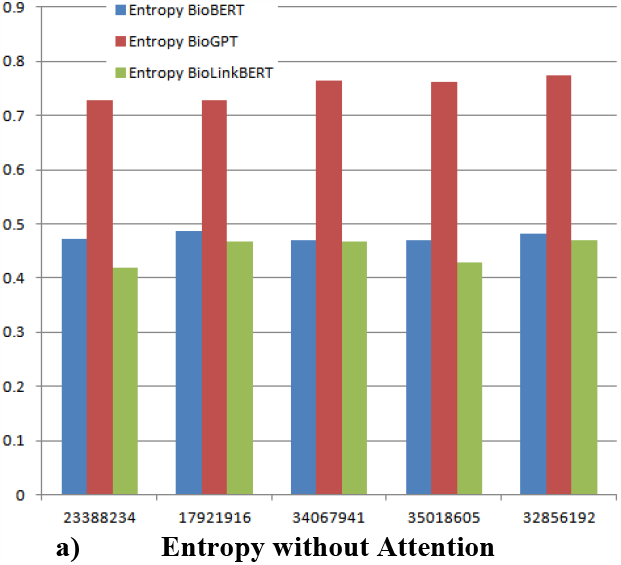
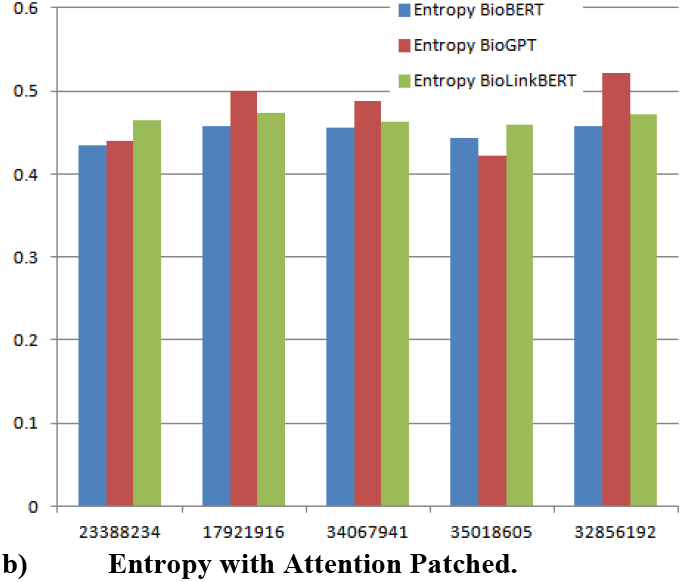
Testing BioBERT, BioGPTand BioLinkBERTwithout and with Attention Patching for Case No. 5 of Cardiothoracic Case in [37].

## VI. Conclusion

Generative models that are based on transformers are designed to understand, generate, and engage in human-like text-based conversation as well as to answer queries by identifying relevant documentations. In evidence based practice this capability is offering significant utility in answering physician queries used for evidence-based medicine. The key features of the transformer models include: highly nuanced language understanding, ability to generate detailed and coherent responses, advanced dialogue management, impressive contextual understanding, identifying relevant materials, and summarization based on extensive trained knowledge base. This helps in providing a high degree of accuracy in interpreting patient information as well as identifying relevant responses to the expected health outcomes. In this paper, we are introducing several attempts to use the generative models for understanding physician PICO questions to predict likely relevant PubMed publications that investigate similar cases. We have introduced two transformer based bootstrapping techniques to identify PubMed relevant articles based on clinical case description (Level 1) and clinical case description with attention patching (Level 2). The transformer models used in the two bootstrapping were the BioBERT and the BioGPT. We verified their performance with BioLinkBERT and find their performance is comparable and better in some cases. Moreover we empowered the attention parching with better similarity function than the Softmax used earlier in Level 1 to using an Entropy function designed as Siamese Neural Network.

## Data Availability

All data produced in the present study are available upon reasonable request to the authors

https://github.com/lukecage0/QL4POMR/tree/main/Sept-Dec

## Acknowledgment

The first and second authors acknowledge the financial support to this research project from MTACS Accelerates Grant (IT22305-2020) and the first author NSERC DDG Grant (DDG-2021-00014).

## Footnotes

2 https://www.webmd.com/

3 https://www.healthtap.com/

4 https://www.healthboards.com/

5 https://www.patientslikeme.com/

6 https://pubmed.ncbi.nlm.nih.gov/

7 https://medlineplus.gov/encyclopedia.html

8 https://www.icliniq.com/

9 https://www.cdc.gov/

10 https://replicate.com/meta/llama-2-13b-chat

11 Thunder Bay Regional Health Science Center (TBRHSC)

## Notes

1* Research supported by NSERC and MITACS.

### Competing Interest Statement

The authors have declared no competing interest.

### Funding Statement

(1) MTACS Accelerates Grant (IT22305-2020) and (2) NSERC DDG Grant (DDG-2021-00014).

